# Altered gene expression profiles impair the nervous system development in individuals with 15q13.3 microdeletion

**DOI:** 10.1101/2022.04.08.22273231

**Authors:** Marek B. Körner, Akhil Velluva, Linnaeus Bundalian, Maximilian Radtke, Chen-Ching Lin, Pia Zacher, Tobias Bartolomaeus, Anna Kirstein, Achmed Mrestani, Nicole Scholz, Konrad Platzer, Anne-Christin Teichmann, Julia Hentschel, Tobias Langenhan, Johannes R. Lemke, Antje Garten, Rami Abou Jamra, Diana Le Duc

## Abstract

**Background:** The 15q13.3 microdeletion has pleiotropic effects ranging from apparently healthy to severely affected individuals. The underlying basis of the variable phenotype remains elusive.

**Methods:** We analyzed gene expression using blood from 3 individuals with 15q13.3 microdeletion and brain cortex tissue from 10 mice Df[h15q13]/+. We assessed differentially expressed genes (DEGs), protein-protein interaction (PPI) functional modules, and gene expression in brain developmental stages.

**Results:** The deleted genes’ haploinsufficiency was not transcriptionally compensated, suggesting a dosage effect may contribute to the pathomechanism. DEGs shared between tested individuals and a corresponding mouse model show a significant overlap including genes involved in monogenic neurodevelopmental disorders. Yet, network-wide dysregulatory effects suggest the phenotype is not caused by a singular critical gene. A significant proportion of blood DEGs, silenced in adult brain, have maximum expression during the prenatal brain development. Based on DEGs and their PPI partners we identified altered functional modules related to developmental processes, including nervous system development.

**Conclusions:** We show that the 15q13.3 microdeletion has a ubiquitous impact on the transcriptome pattern, especially dysregulation of genes involved in brain development. The high phenotypic variability seen in 15q13.3 microdeletion could stem from an increased vulnerability during brain development, instead of a specific pathomechanism.

## Introduction

Individuals with 15q13.3 microdeletion (OMIM #612001) show clinical manifestations ranging from no obvious symptoms to severe intellectual disability, neuropsychiatric disorders, and epilepsy ^1^ (Fig. 1 A, B). The most common 15q13.3 deletion spans 2 Mb and includes eight RefSeq genes (*CHRNA7, FAN1, TRPM1, KLF13, OTUD7A, MTMR10, ARHGAP11B*, and *MIR211*, Fig. 1A) ^2^. Many functional and association studies inquired which gene(s) encompassed by the deleted region could be responsible for the phenotype. However, the results were as variable as the clinical manifestation and different groups proposed multiple candidates (*CHRNA7* ^3,4^, *OTUD7A* ^5,6^, *FAN1* ^7^, *ARHGAP11B* ^8^, *TRPM1* ^9^, *KLF13* ^10^), to explain the symptoms (Fig. 1A). A mouse model of the 15q13.3 microdeletion syndrome (Df[h15q13]/+) shows manifestations similar to affected humans ranging from attention deficits to impaired behavior and disrupted prefrontal cortex processing ^11^. Thus, the microdeletion effect is stable across species and clearly impairs nervous system function.

**Fig. 1.**
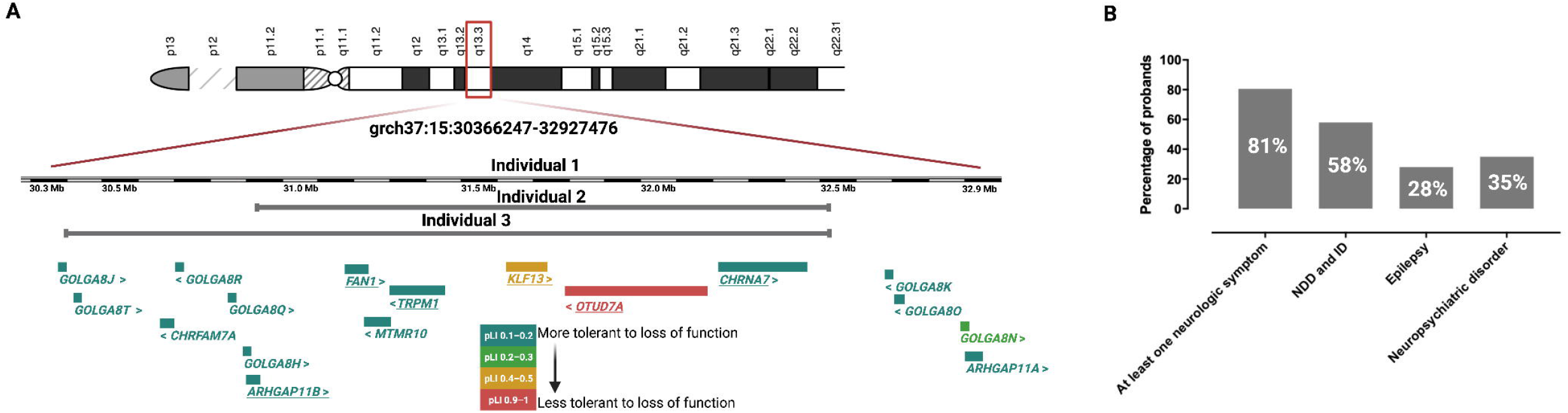
Overview of the 15q13.3 locus and symptoms associated with the microdeletion A. Schematic representation of the 15q13.3 microdeletion region. Protein coding genes within the region are shown beneath chromosome 15. The color legend corresponds to the pLI score as a measure of loss-of-function deleteriousness ^40^. Underlined genes have been considered candidates that are responsible for the observed phenotypes (*CHRNA7* ^3,4^, *OTUD7A* ^5,6^, *FAN1* ^7^, *ARHGAP11B* ^8^, *TRPM1* ^9^, *KLF13* ^10^) B. Individuals with 15q13.3 microdeletion display a heterogenous phenotype which can range from normal development to severe intellectual disability (ID) or neurodevelopmental disorders (NDD). A delineation of the phenotype based on 246 cases revealed predominantly neurologic symptoms of which ID, epilepsy, and neuropsychiatric disorders are most prominent ^2^.

Recently, it was suggested that many disrupted biological pathways such as Wnt signaling or ribosome biogenesis may be involved in the molecular mechanism underlying the disease rather than singular dosage-affected genes ^12^. While Zhang *et al*. used a multiomics approach to identify perturbed biological processes, the multiple employed analyses showed disagreement with respect to the pathomechanism. Also, no shared dysregulated genes were identified between human induced pluripotent stem cells (iPSCs) and mouse cortex ^12^. This could be a result of the *in vitro* setup and neuronal differentiation protocols, which generally impact gene expression profiles ^13^. We thus sought to inquire gene expression profiles in subjects with 15q13.3 microdeletion in a native/*in vivo* state.

One major challenge of transcriptomics in a clinical setting is tissue-specific gene expression ^14,15^ and the fact that most of the times the only accessible tissue to probe is blood ^16^. In our previous work we showed, however, that known genes for neurodevelopmental disorders are not necessarily expressed in the adult brain and that genes which are relevant during embryonic development of the central nervous system can be silenced at a later timepoint ^14^. Thus, although not regarded as a representative tissue, blood transcriptomics has the potential to reveal aspects missed in other tissues or iPSCs.

In the present study, we analyzed the changes in gene expression profiles in the blood of three individuals with heterozygous microdeletion 15q13.3 and intellectual disability associated with epilepsy. We identified a significant overlap (*p*-value = 0.02) of 68 differentially expressed genes (DEGs) between humans and mouse (Df[h15q13]/+) cortex. The gene ontology (GO) category most significantly enriched with DEGs was “nervous system development–GO:0007399” and DEGs in blood, which are not expressed in the adult brain, revealed maximum expression levels in the prenatal stage of brain development. The disrupted gene expression profile could lead to an increased vulnerability in the early stages of nervous system development.

## Materials and Methods

### Ethics approval

This study was approved and monitored by the ethics committee of the University of Leipzig, Germany (224/16-ek and 402/16-ek).

### Chr15q13.2q13.3 microdeletion individuals and mouse model (Df[h15q13]/+)

Three individuals with diagnosed heterozygous 15q13.2q13.3 microdeletions were previously described ^17^. For all individuals we performed Illumina TruSight One Panel and microarray analysis. Patient 1 is a male with a deletion of 2.56 Mbp (15q13.2q13.3(30366247_32927476)x1). The second patient is female and carries a deletion on chromosome 15 of 1.57 Mbp (15q13.2q13.3(30936285_32514341)x1). Patient 3 is a male, has a deletion of 2.14 Mbp on chromosome 15 (15q13.2q13.3(30371774_32514341)x1). He was additionally diagnosed with a maternally inherited splicing-variant in the remaining *TRPM1* allele and a heterozygous *de novo* point mutation in *MITF*. Pathogenic variants in *MITF* gene cause albinism, which was also clinically diagnosed in this individual. His ophthalmological phenotype (severe myopia, astigmatism, and pendular nystagmus) were clinical symptoms of the autosomal recessive TRPM1 phenotype (one allele being deleted as part of the 15q13.3 microdeletion and the other allele carrying the maternally inherited splicing variant). In sum, he suffered from a complex combined phenotype with neurologic symptoms attributed to the 15q13.3 microdeletion. Blood RNA samples were taken from the three individuals (two males and one female, aged 27–63 years) and four control subjects (two males and two females, aged 20–52 years).

To identify molecular changes which are consistent across species we used the data generated by Gordon and colleagues (GSE129891) ^18^. We analyzed transcriptomes from cerebral cortex tissue of mice with heterozygous deletions on mouse chromosome 7qC syntenic to human 15q13.3 ^11,18^.

### RNA extraction and sequencing

RNA was extracted from PAXgene blood samples using PAXgene Blood RNA Kit (Qiagen). RNA sequencing (RNA-seq) libraries were prepared using TruSeq RNA Library Prep Kit v2 (Illumina, San Diego, CA) and sequenced on an Illumina NovaSeq platform with 151 bp paired-end reads.

### Differential Gene Expression (DEG) analysis

RNA-seq reads were mapped to the human genome assembly hg38 with STAR (version 2.6.1d) ^19^. We computed the transcript levels with htseq-count (version 0.6.0) ^20^. From GSE129891 we analyzed read counts of ten wild type and ten (Df[h15q13]/+) mouse cerebral cortex samples. Genes with a sum of less than 10 reads in all samples together were excluded from further analysis. Differential expression of genes was determined with the R package DESeq2 (version 1.30.1) ^21^, which uses the Benjamini-Hochberg method to correct for multiple testing ^22^. Genes were considered to be significantly differentially expressed if *p*-adj < 0.05. To check clustering of RNA-sequencing samples of subjects and controls, a principal component analysis (PCA) was performed with the R package pcaExplorer (version 2.6.0) ^23^. RNA count data were variance stabilized transformed and the 500 most variant genes (top n genes) were selected for computing the principal components.

### Expression of DEGs in different developmental stages

Expression data of DEGs were obtained from PTEE (version 1.1) ^14^ for the adult brain cortex. Genes expressed at a low level in adult brain tissue may be expressed at a higher level in the developing brain and therefore, could still play a significant role in neurodevelopment. Hence, for DEGs expressed <1.5 TPM in adult brain cortex (according to PTEE), expression levels in different developmental stages were obtained from the R package ABAEnrichment (version 1.20.0) ^24^ for the whole brain. We used a Tukey’s HSD test to determine whether this group of DEGs (<1.5 TPM in adult brain cortex) displays a significantly different expression profile between the brain developmental stages.

DEGs which are expressed <1.5 TPM in adult brain cortex but >1.5 RPKM in prenatal stage of the whole brain and reach their maximum of expression in the prenatal stage were selected and further analyzed for GO enrichment.

### DEGs involved in NDDs

A list of genes, which are known to play a role in NDDs, was obtained from PTEE ^14^. DEGs of 15q13.2q13.3 microdeletion patients were compared to the list of NDD genes, to determine DEGs that could contribute to the neurological symptoms observed in those patients. The significance for enrichment of DEGs with NDD genes was calculated using a binomial test in R ^25^.

### GO enrichment

Gene ontology enrichment analysis was performed with the R package GOfuncR (version 1.14.0) for up- and down-regulated DEGs ^26^. GO nodes with a family wise error rate (FWER) <0.05 were considered significantly enriched. To check for unspecific GO enrichment analysis results, the four control subjects were split in two groups and differential gene expression and GO enrichment analyses were performed for those two control groups.

### Identification of activated/inactivated DEG-interacted functional modules

We investigated the activity of DEG-interacted functional modules to elucidate the roles of DEGs in 15q microdeletion. The DEG-interacted network was constructed by the DEGs and their interacting partners in the human protein interaction network (PIN), which was obtained from the InBio Map database ^27^. A DEG-interacted functional module is a subnetwork of the DEG-interacted network formed by genes annotated by the same biological processes. The functional annotations of genes were obtained from GO ^28,29^, and only the annotations supported by experiments were used in this study. Additionally, to ensure the DEGs’ participation and functional association among genes, all the functional modules were required to contain at least one DEG and one interaction. To determine if the member genes of the tested functional module were overrepresented at the top of the entire ranked gene list, we performed the gene set enrichment analysis (GSEA) ^30^ for evaluating each module’s activity and inactivity separately. To assess the activity (inactivity), the entire gene list was ranked downward (upward) by the fold change of genes between the 15q microdeletion and controls. We then calculated the enrichment score (ES) for each functional module by walking down the ranked gene list. The ES of functional module *f* is defined as below:

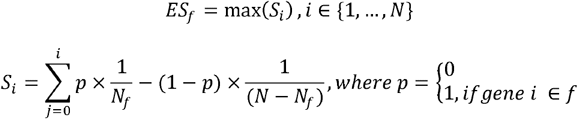

where *S*_*i*_ is the score of gene *i, i* is ordered by fold change, *N*_*f*_ is the number of genes in the tested functional module, *N* is the number of total ranked genes, and *p* is a binary parameter. To estimate the significance of *ES*_*f*_, we produced 1,000 scores *ES*_*rand*_ calculated from 1,000 randomly permutated gene lists. Then, we denoted the standard score *z*, which was defined as below, as the activity or inactivity of functional module *f*.

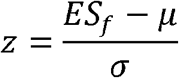

where *μ* and *σ* are respectively the mean and standard deviation of 1,000 *ES*_*rand*_. Finally, the functional modules possessing *z* of activity greater than two and *z* of inactivity less than zero were defined as activated; and the functional modules with *z* of inactivity greater than two and *z* of activity less than zero were defined as inactivated. The discovered activated or inactivated functional modules were further summarized/clustered by the REVIGO ^31^ algorithm with similarity ≥ 0.9 that was calculated from Resnik ^32^ algorithm and visualized using the treemap package ^33^.

To predict key transcription factors and cofactors that drive transcriptomic differences between microdeletion individuals and controls we used Mining Algorithm for GenetIc Controllers (MAGIC), which leverages ENCODE ChIP-seq data to look for statistical enrichment of transcription factors and cofactors in genes and flanking regions ^34^.

## Results

### Transcriptional changes in 15q13.3 microdeletion individuals and Df[h15q13]/+ mice

To study the effects of the 15q13.3 microdeletion on transcriptional regulation, we performed RNA-seq from three individuals carrying a heterozygous 15q13.3 microdeletion (Fig. 1) and four control subjects. Further, to identify robust changes across species and tissues, we analyzed cerebral cortex tissue from ten mice (Df[h15q13]/+) reported by Gordon *et al*. ^18^. While in the 15q13.3 microdeletion subjects we identified 2,334 genes (adjusted *p*-value < 0.05) with altered expression levels compared to controls (Supplementary Table S1), only genes within the deleted region withstood multiple testing correction in the mouse (Supplementary Table S1). This could be related to a difference in synteny between the mouse and human chromosomal regions, to the high interindividual variability of our subjects, or to the generally mild impact on gene expression with genes not reaching the dysregulation threshold necessary to withstand conservative multiple testing correction. The immediate result of applying multiple testing correction is that the probability a true effect may be rejected will increase ^35^. To control for false positives, but also to avoid erroneously rejecting real effects we decided to focus on DEGs shared between human and mouse. We, thus, considered genes with uncorrected *p*-value < 0.05 in the mouse and identified 68 shared genes between the two species and different tissues (Supplementary Table S1). To test whether the number of overlapping genes is higher than expected by chance we performed 100,000 random samplings considering a total of 20,000 genes. This yielded a *p*-value of 0.02 suggesting the overlap is significant. By contrast, when we considered DEGs among controls, only six genes were shared with the (Df[h15q13]/+) mouse model, which is an amount expected to occur by chance (*p*-value=0.94 from 100,000 simulations).

To check whether the gene dosage affects gene expression, we identified genes located in the deleted site, which are expressed in blood and brain cortex (Supplementary Fig. S1). Four genes have an expression higher than 1.5 TPMs in brain cortex ^14^ (*FAN1, MTMR10, KLF13, OTUD7A*) of which *MTMR10* and *KLF13* are also highly expressed in blood (Supplementary Fig. S1). These genes were significantly downregulated in both human and mouse samples (Supplementary Table S1). Moreover, although *FAN1* and *OTUD7A* display low expression levels in blood (Supplementary Fig. S1), they were also significantly differentially expressed in our blood transcriptome analysis (Supplementary Table S1).

We next focused on shared DEGs between 15q13.3 microdeletion individuals and the mouse model. Variants in eight of these genes (*PHIP, KAT6A, VPS13B, GPAA1, CHD7, FIBP, KMT2C, AP1S1*) are known causes for monogenic NDD ^36^. There are six genes which have a gene ontology (GO) annotation related to gene expression (*ZFP57, EDA, KAT6A, CD46, PIK3R3*) and also six genes related to brain development (*CHD7, ITGA4, MYLIP, PAFAH1B3, SIRT2, B4GALT2*). Interestingly, we could also identify components of the major histocompatibility complex, class II to be dysregulated in both blood and brain (Supplementary Table S1), which may reflect a disturbed inflammatory or immune process.

### Molecular pathways affected by transcriptome alterations in 15q13.3 microdeletion

To identify molecular pathways that may be affected by the gene expression profiles we performed GO enrichment analysis followed by protein-protein-interaction (PPI) networks, as previously described ^37,38^. Using the mouse data, we identified general GO categories like cellular components or developmental processes to be enriched with DEGs (Supplementary Table S2). For human subjects, there were two less general GO terms which were most significantly enriched with DEGs: “nervous system development” (GO:0007399, *p*-value after family wise error rate (FWER) multiple correction = 0.036) with 32 associated genes and “DNA binding” (GO:0003677, *p*-value FWER multiple correction = 0.046) with 183 associated genes (Table 1, Supplementary Table S2).

**Table 1.** Enriched overrepresented GO terms in DEGs of 15q13.3 individuals. FWER: family-wise error rate corrected p-value; #genes: number of DEGs involved in the function. For genes included in the nodes refer to Supplementary Table S2.

| GO ID | Ontology | GO term | raw $p$-value | FWER | #genes |
| --- | --- | --- | --- | --- | --- |
| GO:0007399 | biological process | nervous system development | 1.93E-05 | 0.036 | 32 |
| GO:0003677 | molecular function | DNA binding | 0.0001 | 0.046 | 183 |

To better delineate molecular pathways, involved in the copy-number variant (CNV) pathomechanism, we further focused on identifying the functional modules formed by DEGs from human subjects and their PPI partners. This revealed that most nodes clustered in cellular processes like metabolic pathways, signaling, or cellular components (Fig. 2, Supplementary Table S2). Since regulation of gene expression appeared to be perturbed, we tested if the gene expression profile matches dysregulation of one or more transcription factors based on ENCODE Chip-seq data ^34^. This analysis revealed no significant enrichment for genes associated with a known transcription regulator, suggesting that a single gene cannot explain the observed expression profile. Interestingly, inactivated functional modules clusters are mainly involved in immune response and regulation of gene expression (Fig. 2B, Supplementary Table S2). Oligodendrocyte differentiation and development appear to be affected, which together with the “positive regulation of neuron death” (Fig. 2A, B, Supplementary Table S2) could explain the impaired nervous system development. To check whether those molecular pathways are specifically identified in the microdeletion individuals, we analyzed differential gene expression between two control groups. GO terms significantly enriched with DEGs of the control groups were mostly related to immune response, and to a much lesser extent to gene expression regulation (Supplementary Table S2). Thus, the identification of those molecular pathways in the individuals bearing 15q13.3 microdeletion may not be related to the deleted region, but rather to the analyzed tissue. In contrast, we did not identify any GO terms related to nervous system development in the controls. This supports our hypothesis that the effect on pathways related to nervous system development in the affected individuals is a result of the microdeletion.

**Fig. 2.**
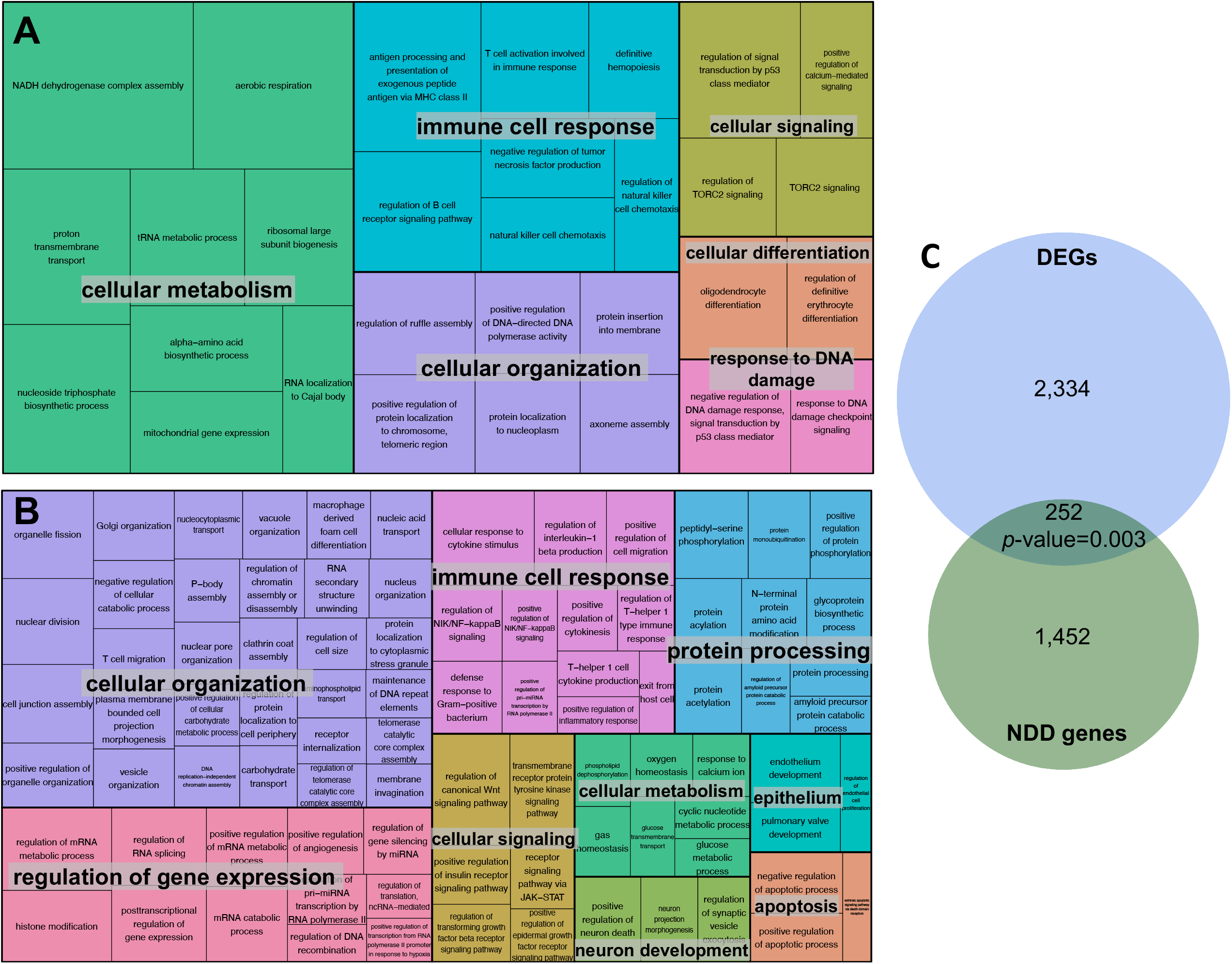
Functional modules representation. A. Activated functional modules clusters in individuals with 15q13.3 microdeletion. Functions that could influence nervous system development are clustered in oligodendrocyte differentiation under the “cellular differentiation” category. The size of the boxes is proportional to the activation level of the module. B. Inactivated functional modules clusters in individuals with 15q13.3 microdeletion. These include processes relevant for neuron development. The size of the boxes is proportional to the inactivation level of the module. C. 252 of the DEGs are related to monogenic neurodevelopmental disorders (NDD). The number of genes is significantly higher than expected by chance (*p*-value binomial test = 0.003).

We showed that affected genes were enriched in pathways related to nervous system development (Table 1) and that PPIs influence apoptosis and neuron death (Fig. 2B). This prompted us to inquire all DEGs that have known Mendelian associations with monogenic NDD. We identified 252 of the DEGs to be related to monogenic intellectual disability (Fig. 2C, Supplementary Table S1). The number of genes is significantly higher than expected by chance (*p*-value binomial test = 0.003), which could suggest an underlying polygenic effect that leads to an increased risk for a neurodevelopmental disorder.

### Dysregulated genes in 15q13.3 microdeletion individuals expressed in the developing brain

Further, we asked whether blood DEGs, which are not expressed in the adult brain cortex, may have been expressed in the developing brain. We identified 358 DEGs, which are expressed in blood but not in the adult brain. We used the ABAEnrichment package in R ^24^ to check the expression levels of these genes during the different stages of brain development (Supplementary Table S3). For 245 of the 358 genes, we could retrieve expression levels from the Allen Brain Atlas. Our analysis revealed that for DEGs, which are silenced in the adult brain, there is a significant enrichment for the ones with a maximum expression level in the developing brain (*p*-value = 0.04, Fig. 3). A GO enrichment analysis of the 53 genes showed that several of these genes are involved in chromosome organization during cell division, but also identified the e.g. *DRAXIN* gene to be dysregulated, which is involved in the development of spinal cord (Supplementary Table S3).

**Fig. 3.**
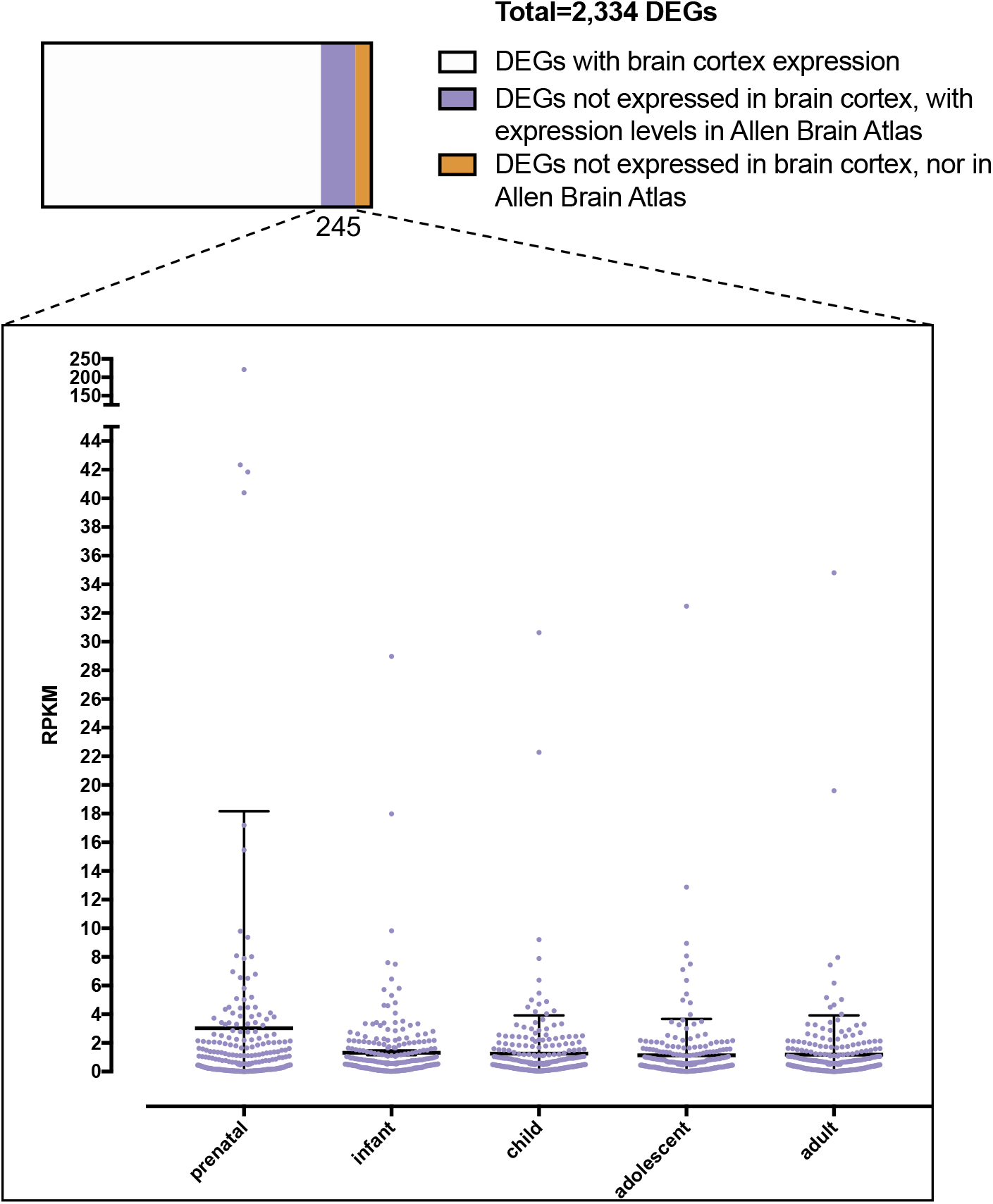
Inquiry of DEGs which are not expressed in the adult brain cortex. Based on the Allen Brain Atlas these genes show a significant enrichment for genes with highest expression level in the prenatal stage (*p*-value adult *vs*. prenatal stage = 0.04).

## Discussion

The 15q13.3 microdeletion is associated with pleiotropic effects and has been described in a wide spectrum of clinical contexts ranging from apparently healthy individuals to some severely affected with ID, epilepsy or even schizophrenia (Fig. 1B) ^2^. It is difficult to dissect the mechanisms contributing to the nervous system developmental disturbance mostly because of the limitations of *in vitro* approaches aiming to reproduce human brain development ^10^. Thus, the etiology of the 15q13.3 microdeletion’s range of hypervariable symptoms remains elusive.

To understand how dysregulation of gene expression contributes to 15q13.3 microdeletion pathomechanisms, we aimed to circumvent artefacts introduced by conventional *in vitro* approaches. Thus, we analyzed transcriptional profiles from three individuals with 15q13.3 microdeletion in an *in vivo* state in blood, which is an easily accessible tissue. To identify genes, which are robustly dysregulated we additionally analyzed brain cortex tissue from a 15q13.3 microdeletion mouse model.

We initially checked for dosage effects of the genes included in the microdeletion (Fig. 1A). This revealed that there are four genes with high expression in brain cortex (Supplementary Fig. S1 ^14^): *FAN1, MTMR10, KLF13, OTUD7A*, all of which showed significant down-regulation in blood (Supplementary Table S1) of our subjects, as well as mouse brain cortex, confirming that a gene dosage effect of the microdeletion contributes to the transcriptional dysregulation. Other genes included in the typical deletion region: *TRPM1, CHRNA7, MIR211, ARHGAP11B*, display low expression levels in brain cortex and blood (Supplementary Fig. S1) and were not significantly differentially expressed in the 15q13.3 microdeletion individuals. While *MIR211* is a microRNA, which was not sequenced probably as a result of the library preparation protocol and *ARHGAP11B* is a human specific gene ^36^, *Trpm1* and *Chrna7* were down-regulated in mouse brain cortex (Supplementary Table S1), further supporting the importance of the haploinsufficiency.

We next focused on dysregulated genes across the two different tissues in the human subjects and the mouse model. One of the shared down-regulated genes is *CHD7*, which is frequently associated with CHARGE syndrome and has been shown to be highly relevant for neuronal differentiation and brain development ^39^. *CDH7*, but also other shared dysregulated genes like *KMT2C* and *KAT6A* are involved in chromatin remodeling and hence in regulation of gene expression. This is in accordance with the findings of Zhang *et al*., who described a global epigenomic reprogramming of iPSCs from 15q13.3 microdeletion individuals ^12^. However, in their approach they were not able to identify driving factors, potentially secondary to the bias induced by *in vitro* cultivation. This could also explain why they do not identify any shared dysregulations with the mouse model and their multiomics approach yielded a rather low correlation level among the multiple analyses.

To identify affected molecular pathways we performed a GO analysis of DEGs. This showed a significant enrichment of DEGs that are involved in DNA binding (Table 1). Moreover, an analysis of functional modules formed by DEGs and their PPI partners confirmed that gene expression regulation is affected (Fig. 2B, Supplementary Table 2). Yet, based on ENCODE data ^34^, we did not identify any transcription factor that could explain the observed transcriptional profile. This is in accordance with the observation of Zhang *et al*. that the disease-relevant impact of the 15q13.3 microdeletion is probably caused by the combinatorial effects of several genes, rather than a single “master” gene. Our analysis showed network-wide dysregulatory effects and explains why knockout models of singular genes encompassed in the deletion could not fully recapitulate the phenotype ^3–10^.

The GO analysis also revealed that DEGs show a significant enrichment in the nervous system development category (Table 1). Indeed, we could show that the set of dysregulated genes contained a significant proportion (*p*-value = 0.003) of genes which have been related to monogenic NDD (Fig. 2C, Supplementary Table S1). The PPI functional module analysis identified more specific developmental processes of the nervous system like oligodendrocyte differentiation, axon and neuron projection development, as well as “positive regulation of neuron death” to be affected (Fig. 2A, B, Supplementary Table S2). This aligns with experimental data from Df[h15q13]/+ mice, which shows that both loss of *OTUD7A* and *CHRNA7* contribute to dendrite outgrowth defects ^3,6^, and also with the recently described involvement of *Klf13* in the development of cortical interneurons. ^10^

To determine whether other DEGs silenced in the adult human brain might have played a role in the nervous system development, we used the Allen Brain Atlas data, which provides brain gene expression data during different developmental stages. ^24^ This showed that a significant number of genes found to be differentially expressed in blood, but silenced in the adult brain had maximum expression levels in the prenatal stage (Fig. 3).

Our data suggests that network-wide dysregulatory effects contribute to 15q13.3 microdeletion pathomechanisms. There are several lines of evidence that indicate a disturbed nervous system development, suggesting the severity of 15q13.3 microdeletion individuals’ symptoms is probably determined in the early embryonic stages. The identification of dysregulated genes clustering in inflammatory and immune pathways may be related to the analyzed tissue, namely blood. However, since we identified components of the major histocompatibility complex, class II to be dysregulated in the mouse brain, we cannot rule out that immune insults could contribute to the increased vulnerability of 15q13.3 microdeletion-bearing offspring.

A major limitation of our study is the small cohort, in which individual-characteristic gene expression levels, which were not caused by the microdeletion, can have a big impact on the analysis. We attempted to circumvent this by comparing our data to the mouse model. However, a larger cohort and potentially the analysis of an additional tissue like skin, could further refine the analysis and unveil which gene network is mostly responsible for the pathomechanism. This is crucial for directing future efforts to minimize the severity of the phenotype.

## Data Availability

RNA sequencing reads and expression profiles have been submitted to the Gene Expression Omnibus (http://www.ncbi.nlm.nih.gov/geo/) under accession number GSE197903. The code used for analyzing data has been deposited under https://github.com/akhilvelluva/15q13.3.

https://www.ncbi.nlm.nih.gov/geo/query/acc.cgi?acc=GSE129891

## Acknowledgements

We are grateful that our patients and their families agreed to participate in this study. We thank Sandra Schinkel, Kathleen Lehmann, and Sophie Behrendt for their great technical assistance and Rigo Schulz for server support. We are grateful to Torsten Schöneberg for his input and help to draw Fig. 1.

## Author contributions

MK performed gene expression analyses, contributed to the design of the study and writing of the manuscript. AV and LB supported bioinformatic analyses and performed GO enrichment analyses. MR performed wet lab work and contributed to the design of the study. CCL performed PPI analyses and contributed to manuscript writing. PZ recruited patients, performed phenotyping, and coordinated the genetic diagnosis. TB, AT, KP, and JH performed genetic diagnosis and contributed to the writing of the manuscript. AK, AM, NS, TL, KP, JRL, and AG contributed to analysis design, data interpretation, and writing of the manuscript. RAJ and DLD designed the study, coordinated the contact to patients, contributed to genetic diagnosis, gene expression analysis, and writing of the manuscript.

---

**Supplementary Fig. S1.**
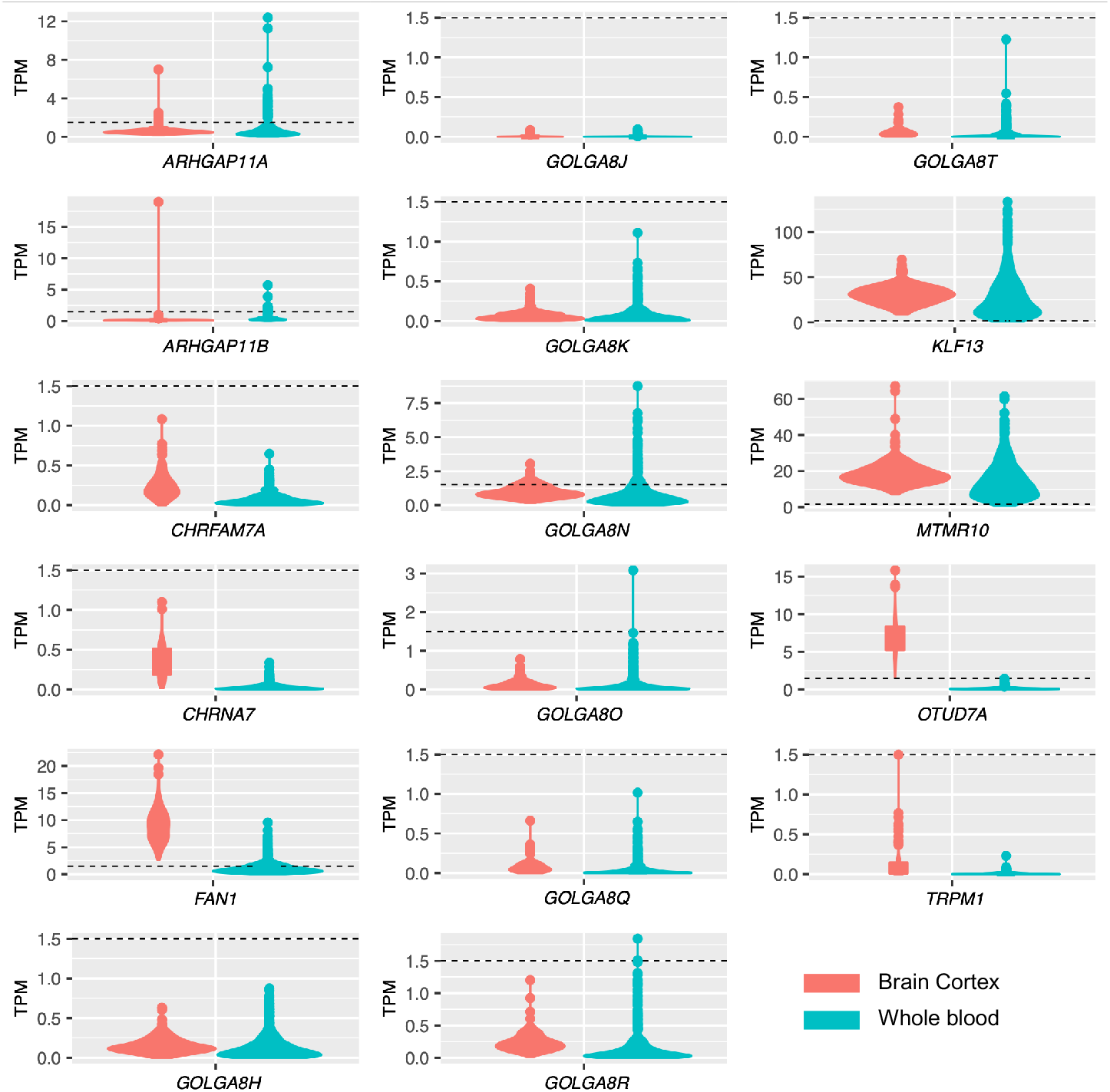
Expression levels of genes located in the 15q13.3 microdeletion region. TPM (transcript per kilobase million mapped reads) values of gene expression levels are depicted for brain cortex tissue and whole blood. The dashed lines represent 1.5 TPM. Expression values were obtained from PTEE (https://bioinf.eva.mpg.de/PTEE/).

